# Covid-19 Related Generalized Anxiety Disorder Among Health Care Workers In A Hospital In The Greater Accra Region

**DOI:** 10.1101/2023.10.16.23297097

**Authors:** Ohakpougwu Chukwuebuka Emmanuel, Wendy M. Bebobru

**Affiliations:** Accra Psychiatric Hospital; Family Health University College; Nyaho Medical Centre

## Abstract

**Background:** The emergence of Coronavirus disease 2019 (COVID-19) an infectious disease caused by the newly discovered severe acute respiratory syndrome coronavirus 2 (SARS CoV-2) has caused a lot of harm to humanity. Healthcare workers who are the leading the charge in the fight against the virus can experience mental health challenges with anxiety being an important illness. Anxiety can become morbid quickly and ultimately affect function, hence the need to study its prevalence among HCWs, since they are a high-risk population. Studies across various regions worldwide reported elevated levels of anxiety amongst HCWs during the SARS, Ebola, and H1N1 pandemics. Nevertheless, Generalized Anxiety Disorder (GAD), an easily measured and ubiquitous member of the family of anxiety disorders has hardly been researched. However, new studies in Togo, China, India and Mexico have reported elevated levels of GAD in HCWs during the COVID-19 pandemic. Given the complexities surrounding mental health care in Ghana, and Africa as a whole it would be expedient to uncover the prevalence of GAD among HCWs during the pandemic. Hence, a study at Family Health Hospital will provide information about the prevalence of COVID-19 related GAD among Health care workers representative of Ghana.

**Aim:** The aim of this study was to establish the prevalence of COVID-19 related GAD amongst healthcare workers, in a tertiary hospital in Accra.

**Methods:** A descriptive cross-sectional study design using a self-administered questionnaire was employed. Nine-two (92) HCWs in the study area were sampled. A consecutive sampling technique was used to select the respondents for the study. The study was analyzed using SPSS version 25. The results were presented in summary tables and analyzed using frequencies and percentages. Chi square test performed on categorical data to test association between selected variables and their outcome with COVID-19 related GAD.

**Results:** The GAD level among nurses was 55.4%, and for doctors it was 30.4%. The GAD level among medical laboratory technicians and pharmacists were 7.6% and 6.5% respectively. Furthermore, being age 50-69 years was a significant risk factor for developing GAD during the COVID-19 pandemic in Ghana. Female HCWs were more likely to experience GAD. However, only 13.1% of the HCWs were considered to have Corona phobia. Perception of workplace as being high risk was positively correlated with mild to moderate forms of anxiety. However, perception of organizational support as being guaranteed in case one succumbed to the virus and confidence in PPE availability was not reported to be strong protective factors against GAD among HCWs.

**Conclusion:** COVID-19 related GAD is a challenge amongst HCWs especially nurses in FHH. The management of the FHH should set up certain services such as psychological help lines, peer support programs as well as run a sensitization campaign to cater for the wellbeing of doctors as well as encourage mental health seeking behavior.

## INTRODUCTION

### 1.1 Background

The emergence of COVID-19 an infectious disease caused by the newly discovered severe acute respiratory syndrome coronavirus 2 (SARS CoV-2) has caused a lot of harm to humanity. COVID- 19 was first reported from Wuhan, China on the 31^st^ of December 2019 as a cause of pneumonia and since then has spread to many countries around the globe causing the World Health Organization (WHO) to declare it a pandemic on the 11^th^ of March 2020 (WHO, 2020). As at July 29^th^ 2020, there have been over 16 million confirmed cases and 655,112 deaths (Worldmeters.info, 2020). According to the Africa Centre for Disease Control (Africa CDC) as at July 29^th^ 2020, South Africa had the highest number of confirmed cases (459,761), followed by Egypt, Nigeria, and Ghana (Africa CDC, 2020). As at the 29^th^ of July, Ghana had 34,406 confirmed cases and 168 deaths. On the 18^th^ of July 2020 Cable News Network reported that 2000 HCWs had been infected since March when the first case in Ghana was reported (CNN, 2020). The burden of COVID-19 is not just limited to physical health but it also affects mental health (Galea et al, 2020). According to previous studies from SARS and Ebola pandemics the onset of a sudden and immediately life threatening illness could lead to extraordinary amounts of pressure on healthcare workers (Liu et al, 2012). Increased workload, physical exhaustion, inadequate personal protective equipment, nosocomial transmission and the need to make ethically difficult decisions on rationing care may have dramatic effects on their physical and mental wellbeing (Pappa et al, 2020). Their resilience can be further compromised by isolation and loss of social support, risk of infection of friends and relatives as well as drastic, often unsettling ways of working (Pappa et al, 2020). HCWs are, therefore, especially vulnerable to mental health problems, including fear, anxiety, depression, and insomnia (Lung et al., 2009; Wu et al., 2009). Anxiety can be a physical and/or a psychological reaction to stress and can in fact be beneficial to help us anticipate potential dangers and prepare for them. Physically, it can present as increased sweating, palpitations, tachypnea, and dizziness amongst other symptoms. Psychologically, it presents as “feeling worried all the time”, “feeling tired”, poor sleep (Royal College of Psychiatry, 2015). In general, for a person to be diagnosed with anxiety disorder the anxiety must fulfill the following criteria: be out of proportion to the situation or age inappropriate, hinder your ability to function normally, the severity of distress caused by the symptoms of anxiety (American Psychiatric Association, 2017). Thus an anxiety disorder may make people feel anxious most of the time or for brief intense episodes, which may occur for no apparent reason (Rector et al, 2008) According to the American Psychiatric Association (APA, 2017) Anxiety disorders are the commonest mental disorders and affect nearly 30 percent of adults at some point in their lives. The WHO reports that as at five years ago, an estimated 3.6%(264 million) of the world’s population was living with anxiety disorders (WHO, 2015). This total estimate is higher by 14.9 percent than it was in 2005, and ten percent of the aforementioned estimated population resided in Africa (WHO, 2015). There is no singular cause of anxiety disorders rather it is caused by a myriad of factors such as stressful or traumatic life events, family history of anxiety disorders, childhood development issues, alcohol, medications, or illicit substances, and medical or psychiatric problems (Rector et al, 2008). Anxiety disorders are grouped into 6, which include generalized anxiety disorder, panic disorder, phobias, agoraphobia, social anxiety disorder, and separation anxiety disorder.

This study is focused on Generalized Anxiety Disorder (GAD). Generalized anxiety disorder involves “excessive anxiety and worry, occurring more days than not for a period of at least six months, about a number of events or activities”. It is characterized by difficulty to control worry” (APA, 2000, p.472-6). Furthermore according to the Diagnostic and Statistical Manual -Five (DSM 5, 2013), the anxiety must be associated with any three of the following: restlessness, easy fatiguability, irritability, muscle tension, and sleep disturbance. The lifetime prevalence of GAD is 9% and its prevalence in 12-months is 3.1%(DSM-5, 2013). Its median age of onset is 31, and life stressors are a major precipitant. GAD affects daily life and interferes with ability to take appropriate decisions. Workplace impairment studies have reported a reduction in overall work productivity in the range of 11-49% in more than 23% percent of individuals with pure GAD, and more than 50% in 20% of individuals with comorbid Major depressive disorder (Wittchen et al, 2000). Given that most people with GAD are employed, this forebodes a substantial cost to employers and society. The crucial role of HCWs during a pandemic as front liners is vital and massive, making them susceptible to anxiety and stress. In the early stages of the pandemic, infected HCWs accounted for 29% of all hospitalized COVID-19 patients. Therefore they are subject to complex emotional reactions and psychological distress, which can impair their attention, cognitive functioning, and clinical decision-making (Leblanc V, 2009; Pangioti et al, 2018). In the fight against COVID, “time is breath”. Thus if HCWs have impaired ability to take critical decisions for management or promptly identify symptom clusters then more life will be lost and there is greater chance of infection transmission. Prolonged unrecognized anxiety could lead to major psychiatric morbidity, exhaustion, and resignations from duties due to moral injury (Alharty et al, 2017). Furthermore early recognition of GAD will prevent its progression to depression and its attendant economic loss due to the burden of disease. Thus it is the goal of this study to determine the prevalence of GAD among HCWs, who are the heroes of the fight against COVID-19 in order to suggest psychological interventions that will benefit them as part of a robust psychological support program. Unfortunately, there was only article (from Togo) tackling Anxiety disorders in HCWs on the African continent. This study showed increased levels of anxiety amongst doctors and nurses. Nevertheless, several studies from outside the continent have clearly shown elevated levels of anxiety in health care workers (Paybast et al, 2020). However, there were no studies focused specifically on GAD.

### 1.2 Problem Statement

The emergence of COVID-19 has brought about untold suffering to the world ranging from economic, physical and psychological. Various authors have written extensively on the psychological impact of COVID-19 on the general population thus leading to increased levels of depression, anxiety, and suicides (Li et al, 2020; Qiu et al, 2020; Rubin et al, 2020; Wang et al, 2020). Furthermore, there has been the aggravation of already existing psychopathology in patients with mental health challenges as result of this pandemic (Dubey et al, 2020). Health workers are particularly at risk of elevated levels of anxiety and stress due to the nature of their service. Previous studies on the mental health of health care workers during the SARS pandemic revealed elevated levels of stress depression and anxiety especially amongst nurses followed by doctors (Liu et al, 2012; Su et al, 2005). Anxiety, when excessive can negatively impact an individual’s decision-making abilities. This impairment of function and critical thinking is particularly costly in healthcare because human life is at great risk all the time. Furthermore, with pandemics there are acute traumatic patient presentations in great numbers and can lead to long-term psychological consequences (Lin et al, 2007). This can negatively influence a health care workers behavior subsequently. Again, there is also the concern of contracting the virus or transmitting the infection to family and loved ones (Nickell et al, 2005). Anxiety when excessive can present as Generalized Anxiety Disorder (GAD). Thus, it will be expedient to determine the occurrence of GAD amongst healthcare workers especially in Africa where mental health services are grossly underfunded and stigma is also a big factor compounding health-seeking behavior (Bazghina-werq et al, 2020).

### 1.3 Justification

The negative impact of GAD on the mental wellbeing of doctors cannot be over-emphasized. Thus, there is a need to heal and protect our healers (Kalaichandran, 2020). The results generated from this study will justify the need for a paradigm shift in the COVID-19 protocols at various hospitals in the country, as doctors will be encouraged to seek psychological help. Again, this study will encourage the development of hotlines and other mental health and psychosocial services (MHPSS) for HCWs to reach out for help once they start to experience my symptoms positive for GAD and by extension other mental health challenges. This study can influence the drafting of a hospital policy that checks in on doctor’s bi-weekly during this pandemic by getting them to participate in quick surveys to assess their mental health status. All the aforementioned policy shifts will cater to the wellbeing of doctors and nurses to ensure that no lives are lost due to mental health challenges during this pandemic. With the justifications stated above the following research questions have been formulated to address the research topic

### 1.4 Research Questions

1. What factors predispose HCWs to developing GAD during the COVID-19 pandemic?
2. What is the prevalence of GAD among HCWs during the COVID-19 pandemic?
3. Which group of HCWs is more likely to develop GAD?

### 1.5 Aims

The aim of this study is to establish the prevalence of COVID-19 related GAD amongst healthcare workers, in a tertiary hospital in Accra.

### 1.6 Objectives

1. To determine the prevalence of GAD amongst HCWs during the COVID-19 pandemic.
2. To establish the relationship between perceived high-risk work station/unit and GAD during the COVID-19 pandemic.
3. To identify higher levels of GAD among a particular group of HCWs during the COVID-19 pandemic.

### 1.7 Research Hypothesis

1. Age and CAS score

H_0_ : There is no relationship between age and coronavirus anxiety score (CAS) H_1_: There is a relationship between age and CAS score

2. Age and GAD score

H_0_ : There is no relationship between age and Generalized Anxiety Disorder (GAD) score H_1_: There is a relationship between age and s Generalized Anxiety Disorder (GAD) score

3. GAD and the various professions

H_0_ : There is no relationship between GAD score and the branch of the health care in which a HCW specializes .

H_1_: There is a relationship between GAD score and the branch of the health care in which a HCW specializes .

4. GAD and sex/gender

H_0_ : There is no relationship between GAD score and the gender of the HCW H_1_: There is a relationship between GAD score and the gender of the HCW .

5. GAD and workplace risk

H_0_ : There is no relationship between GAD score and the perception by the HCW that a workstation is high risk.

H_1_: There is a relationship between GAD score and the perception by the HCW that a workstation is high risk .

6. GAD and previous psychiatric diagnosis

H_0_ : There is no relationship between GAD score and previous psychiatric diagnosis of the HCW.

H_1_: There is a relationship between GAD score and previous psychiatric diagnosis of the HCW.

7. CAS and fear of contracting covid19 at work

H_0_ : There is no relationship between CAS score and fear of contracting covid19 at work. H_1_: There is a relationship between CAS score and fear of contracting covid19 at work.

8. CAS and occupational support

H_0_ : There is no relationship between CAS score and occupational support at work. H_1_: There is a relationship between CAS score and occupational support at work.

## LITERATURE REVIEW

The burden of COVID-19 is not just limited to physical health but it also affects mental health. According to previous studies from SARS and Ebola pandemics the onset of a sudden and immediately life threatening illness could lead to extraordinary amounts of pressure on healthcare workers (Liu et al, 2012).

### 2.1 Definitions Anxiety

It can be a physical and/or a psychological reaction to stress and can in fact be beneficial to help us anticipate potential dangers and prepare for them. Physically, it can present as increased sweating, palpitations, tachypnea, and dizziness amongst other symptoms. Psychologically, it presents as “feeling worried all the time”, “feeling tired”, poor sleep (RPsych, 2015). In general, for a person to be diagnosed with anxiety disorder the anxiety must fulfill the following criteria: be out of proportion to the situation or age inappropriate, hinder your ability to function normally, the severity of distress caused by the symptoms of anxiety (APA, 2017).

#### Generalized anxiety disorder

This involves “excessive anxiety and worry, occurring more days than not for a period of at least six months, about a number of events or activities”. It is characterized by difficulty to control worry” (APA, 2000, p.472). Furthermore according to the Diagnostic and Statistical Manual (DSM- 5, 2013), the anxiety must be associated with any three of the following: restlessness, easy fatiguability, irritability, muscle tension, and sleep disturbance.

#### Corona phobia

Simply put, corona phobia is fear and anxiety about COVID-19. Although a relatively new construct, research has begun to show that it is indeed a relevant entity and individuals considered to have corona phobia experience a set of unpleasant physiological symptoms that are triggered by thoughts or information about this infectious disease(Lee et al, 2020). Again, corona phobia has been shown to be strongly associated with depression, GAD, hopelessness, suicidal ideation, and functional impairment (Lee et al, 2020).

### 2.2 Risk Factors

#### Socio-demographic factors

Being female, and being a young medical staff (< or = 30 yrs) have both been shown to be important socio-demographic factors in the development of anxiety among health workers (Lai et al, 2020, Liang et al, 2020).

#### Personality and coping strategies

Studies in Mainland China and Taiwan have revealed that neuroticism as a personality trait and maladaptive coping are positively correlated with decreased mental health outcomes (Lu et al, 2006; Lung et al, 2009). Adaptive coping measures can lead to low psychiatric morbidity (Phua et al, 2005).

#### Past psychiatric history and a history of traumatic life events

Some retrospective studies in Toronto and Taiwan among HCWs during the SARS pandemic revealed that having a past history of mental illness increased ones risk of experiencing anxiety and other psychiatric symptoms (Lancee et al, 2008; Su et al, 2007). Furthermore reporting traumatic life events was also found to contribute to the onset of psychiatric symptoms in HCWs during a pandemic (Lung et al, 2009; Wu et al, 2009).

### 2.3 Work Related Factors Anxiety about infection at work

Physicians were found to be less anxious about infection compared with other HCWs, according to studies from Saudi Arabia, Japan and China (Alsubaie et al, 2019; Matsuishi et al, 2012). In addition, nurses experienced more psychological symptoms, including anxiety (Lai et al, 2020; Wong et al, 2005).

#### Level of exposure/ low risk or high risk ward

During the MERS-CoV and SARS outbreaks, HCWs in units at high risk of infection present with more severe mental health outcomes compared with HCWs in units with low risk of infection (Bukhari et al, 2016). Also, it was shown that direct contact with affected patients is a significant risk factor for poor mental health outcomes among (Chong et al, 2004; Grace et al, 2005)

#### Protective factors

Resilience indicators (vigor and hardiness) and self-efficacy were reported to improve mental health outcomes among nurses during the SARS pandemic (Ho et al, 2005; Marjanovic et al, 2007; Park et al, 2018).

#### Organizational variables

Two Canadian studies report organizational support as a protective factor among nurses during the SARS outbreak (Fiksenbaum et al, 2006; Lancee et al, 2008). Again, perceived adequacy of training was also recognized to be an important protective factor (Maunder et al, 2006, Tang et al, 2017).

#### Confidence in PPE and protective measures

This appeared to improve mental health outcomes with regards to stress and burn out (Marjanovic et al, 2007; Preti et al, 2020) but not necessarily the general mental state(Styra et al, 2008)

### 2.4 Anxiety amongst health workers during the SARS pandemic

During the SARS pandemic in 2003, anxiety was prevalent among health care workers in Hong Kong. Anxiety was more prevalent in nurses (52.0%), than doctors (47.8%) during that pandemic (Poon et al, 2004). A study in Taiwan also showed increased levels of anxiety alongside depression during the SARS pandemic (Cheng-Sheng et al, 2005)

### 2.5 Anxiety amongst health workers during the Ebola pandemic

A study of anxiety levels amongst health professionals at an Ebola treatment unit in Liberia revealed anxiety especially among male staff and staff responsible for cleaning and disinfection (Li Li et al, 2020). Increased levels of anxiety among HCWs were also confirmed to be a mental health challenge in Sierra Leone during the Ebola pandemic (Ji et al, 2017).

### 2.6 Anxiety amongst health workers during the A/H1N1 pandemic

In a general hospital in Japan during the outbreak of the H1N1 flu, health workers in high- risk environments had increased anxiety compared to those working in low risk environments. Nurses appeared to have higher anxiety levels during the H1N1 pandemic compared to other groups of health care workers (Matsuishi et al, 2012). A study at a University General Hospital in Greece reported that more than half (56.7%)of the HCWs had moderately high anxiety about the pandemic (Goulia et al, 2010).

### 2.7 Anxiety amongst health workers during the COVID-19 pandemic

Health care workers were shown to be at a great risk of developing symptoms of anxiety disorder, and other psychiatric conditions like depression and insomnia in China. Also, being female and being in wards where there is greater chance for coming in contact with COVID-19 patients were positively associated with anxiety, insomnia, depression, and obsessive compulsive disorder (Zhang et al, 2020). A study narrated by Chakraborty (2020) of medical staff at a tertiary hospital corroborated the finding in the earlier study. Here, being female and working as a nurse was positively associated with increased anxiety. Multi-sectional studies at various hospitals in Iran, and India support the aforementioned findings and also posit that being between the ages of 30 to 40 was significantly related to increased anxiety (Wilson et al, 2020; Zandifar et al, 2020). A narrative review by Thapa et al (2020), revealed that anxiety (44.6%) polled high among health workers, only second to depression (50.4%). However, an observational study by Tan (2020) in Singapore negated the aforementioned statistics. The study of HCWs in Singapore posits that anxiety disorder was more common than any other mental health disorder during the COVID-19 pandemic. This study also showed a higher prevalence of anxiety among non-medical health workers.

With regards to level of training and its influence on experiencing GAD, first year residents in China were shown to have significant levels of depression and anxiety (Li et al, 2020). In addressing the sources of anxiety among HCWs, a report by Shanafelt et al (2020) in the United States, informed that HCWs had eight great sources of anxiety namely: access to appropriate PPE, fear of being exposed to COVID-19 at work and taking it home to family, not having rapid access to testing if they suspect they have symptoms, and uncertainty about organizational support if they get infected, amongst other fears. Being quarantined was a significant cause of anxiety among HCWs, according to a Vietnamese study (Do Duy et al, 2020). HCWs were found to have a significant anxiety rate (62.9%) in Togo. Where further grouped, they were found to have 25.8%, 22.6%, and 14.5% rates for mild, moderate and severe anxiety respectively (Kounou et al, 2020). This was high despite the relatively low infection rates in Togo. Kounou et al (2020) opined that these high rates could be accounted for by the poor quality of health care and lack of adequate PPE to deal with the COVID-19 pandemic in Togo. In addition, Agberotimi et al (2020) after their study in Nigeria reported that health care workers experienced significantly more anxiety (58.4%), compared to the general population (49.6%). Unfortunately, those were the only studies from Sub Saharan Africa and even Africa as a continent that were related to the research topic. The results of

the Togolese study were similar to those from Saudi Arabia where the majority of doctors had mild anxiety (Temsah et al, 2020). A study of 231 HCWs in Mexico City using the Coronavirus Anxiety Scale (CAS) revealed that 30.3% of the participants were considered to have corona phobia having scored > or = 9 on the scale (Ignacio et al, 2020). Furthermore, Ignacio et al (2020) reported that working in high exposure settings, and being female were positively correlated with high CAS score and higher levels of GAD. Almost all articles reviewed, studied mental health challenges/ disorders in HCWs during the pandemic as a broad topic. Thus, there was a paucity of articles that specifically studied GAD in HCWs during the COVID-19 pandemic. This is a lacuna in our knowledge that this present study intends to fill. This is especially important given the pace at which GAD can become morbid.

## METHODOLOGY

### 3.0 Introduction

This chapter describes the various methods and techniques that will be used for this study. It includes description of the study design, study site, the study population, sample size, sampling method, data collection instruments and methods, the dependent and independent variables and how the data analysis was done after data collection.

### 3.1 Study Design

The research design for the study will be a descriptive cross-sectional study, using questionnaires to assess the experience of symptoms of overt anxiety amongst HCWs at FHH.

### 3.2 Study Area

The study will be conducted at Family Health Hospital (FHH). It is a 24hour private hospital established in 1993 under the Companies Code 1963, Act 179 to provide diagnostic, laboratory and medical services to both corporate and individual clients. It started operations from a rented premise at Zoti Road, Latebiokoshie but now operates from its own premises at Teshie, opposite Kofi Annan International Peace Keeping Training Centre (KAIPTC) within Ledzokuku Krowor Municipality in the Greater-Accra Region of Ghana.

Family Health Hospital has a 70-bed capacity and sees about 4000 patients per month. The hospital has staff strength of about 140, of which 120 are health workers. The hospital offers Medical, Pediatric, Surgical, Obstetrics and Gynecology, Dermatology, ENT and Urology specialties. There also exists an In-vitro Fertility Centre, an eye clinic and an Accident and Emergency Centre. A Public Health Unit is also present. (fhu.edu.gh/hospital/, 2020)

### 3.3 Study Population

The study population will be made up of a cross section of HCWs at FHH ranging from, doctors, nurses, to pharmacists.

### 3.4 Inclusion Criteria

All HCWs who are considered Health Care Professionals (HCPs) like doctors and nurses and Health Care Associates like medical laboratory technician and pharmacists under the umbrella of health service providers. Strictly speaking HCWs are all people engaged in actions whose primary intent is to promote health (WHO, 2006). However for the purpose of this study all those who met the criteria of being professionals or associates were included.

### 3.5 Exclusion Criteria

HCWs who could not participate in the study because of their tight schedule and those who were not present when the questionnaires were administered were excluded.

### 3.6 Sample Size

The target population was selected from healthcare workers at FHH. Sample size was calculated using The Yamane Equation where “*n”* is the sample size, “*N”* is the population size of HCWs at 37 Military Hospital

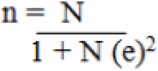

N= 120/ [1+120(0.05) ^2] = 92

N (120) and “*e”* stands for precision value of 0.05%, using 95% confidence. This gave a sample size of 92.

GAD will be classified based on the GAD-7 Score as follows 0-4 (minimal anxiety), 5-9 (mild anxiety), 10-14 (moderate anxiety), 15-21 (severe anxiety). The CAS score will also be used as follows: a score > or = 9 indicates probable dysfunction that warrants further assessment.

### 3.7 Sampling Method

Consecutive sampling method will be utilized to obtain the target sample size. Here every HCWs meeting the criteria of inclusion are added or selected to participate in the study until the required sample size is achieved.

### 3.8 Data collection process

A self-administered questionnaire was designed. This questionnaire was made up of 4 sections. Section A covered informed consent whilst section B handled socio-demographics like age, sex, marital status, profession and possible influences on anxiety such as “considering working environment as high risk or low risk” and hospital department. Section C comprised of the 7 items Generalized Anxiety Disorder score (GAD-7). Section D comprised of the Coronavirus Anxiety Scale (CAS), a novel way of clinically assessing anxiety. The CAS is a 5 item mental health-screening tool to help identify possible dysfunctional anxiety due to COVID-19. The CAS has exhibited good diagnostic properties and demonstrates 90% sensitivity and 85% specificity for dysfunctional anxiety (Lee et al.,2020).

### 3.9 Data Management And Statistics

Data will be collected and captured electronically by the principal researcher under the supervision of the study supervisor and then subsequently entered into Microsoft Excel ® and analysed using IBM SPSS version 25. The results would be presented in summary tables and charts and analyzed using frequencies and percentages. Associations between variables were determined using a non-parallel test at p-values than 0.05, which were considered statistically significant.

#### 3.9.1 Quality Assurance Pretesting the research tool

The research tool was being pretested on a few medical staff at FHH. This informed some modifications that were made to the questionnaire.

#### 3.9.4Ethical consideration

Ethical approval will be obtained from the Community Health Department Review Committee of the University of Ghana Medical School, and the Medical Director of FHH.

## RESULTS

### 4.0 Introduction

This chapter focuses on a detailed analysis of the primary data obtained during the study assessing the prevalence of COVID-19 related GAD among HCWs, as well as contributing factors. The Statistical Package for Social Science (SPSS) version 25 and Microsoft Excel version 15 were tools used for the data analysis. Furthermore, the results are demonstrated via frequency tables, bar and pie charts. A Chi-square test was also performed.

### 4.1 Socio-demographics

#### 4.1.1 Gender of Respondents

In the study male and female personnel across four medical fields chosen from the sample population responded to the items. The data revealed that majority of the respondents were nurses, which represent 55.5% of the sample population of which 53.3% were female and 2.2% were male. Of the 28 respondents who identified as doctors, 18 (19.6%) of the sample population and 10 (10.9%) were females. Of the 6 respondents who identified as pharmacists, 3 were male and 3 were female which represents 3.3% each of the sample population. However, the data show that all the 7() respondents who identified as med. Lab scientists were male. The result was further presented in a bar chart to aid visual clarity.

**Table 1:**
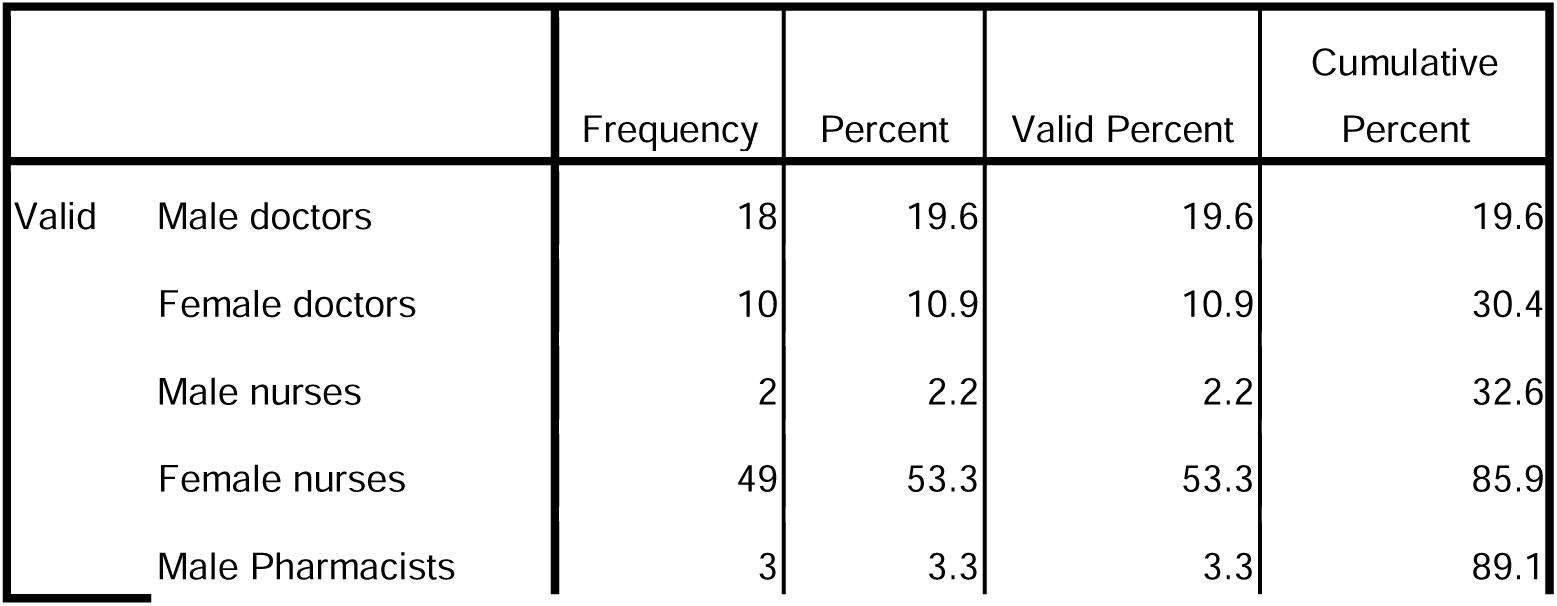
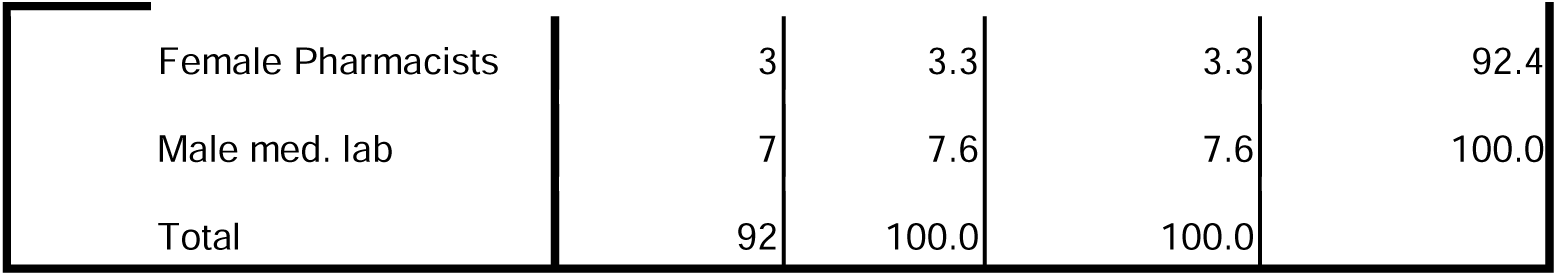
Gender of respondents across professions.

|  |  | Frequency | Percent | Valid Percent | Cumulative Percent |
| --- | --- | --- | --- | --- | --- |
| Valid | Male doctors | 18 | 19.6 | 19.6 | 19.6 |
|  | Female doctors | 10 | 10.9 | 10.9 | 30.4 |
|  | Male nurses | 2 | 2.2 | 2.2 | 32.6 |
|  | Female nurses | 49 | 53.3 | 53.3 | 85.9 |
|  | Male Pharmacists | 3 | 3.3 | 3.3 | 89.1 |

|  |  |  |  |  |
| --- | --- | --- | --- | --- |
| Female Pharmacists | 3 | 3.3 | 3.3 | 92.4 |
| Male med. lab | 7 | 7.6 | 7.6 | 100.0 |
| Total | 92 | 100.0 | 100.0 |  |

#### 4.1.2 Age and Coronavirus Anxiety Score (CAS)

**Table 2:** Age Distribution of Sample * Distribution CAS Cross tabulation.

| Independent Variable | CAS score | | $\chi^2$<br>(Pearson Chi-square) | p-value |
| --- | --- | --- | --- | --- |
| Age (Years) | <9 | > or =9 | 83.73 | 0.000 |
| 21-29 | 24 | 0 |  |  |
| 30-39 | 41 | 0 |  |  |
| 40-49 | 15 | 1 |  |  |
| 50-59 | 0 | 5 |  |  |
| 60-69 | 0 | 6 |  |  |
|  | 80 | 12 |  |  |

The table above presents the cross tabulation of coronavirus anxiety score (CAS) scores and age distribution of respondents. It was observed that all 24 respondents who identify with a less severe CAS (<9) were between 21-29 years of age. Similarly, 41 respondents who suffer low CAS (<9) were between the 30-39 years of age. Likewise, 15 respondents who suffered less CAS (<9) was between the ages of 40-49, while a single respondent with severe CAS (>=9) were between the ages of 40-49. Furthermore, respondents between the ages of 50-59 and 60-69 who suffered severe CAS (>=9) were 5 and 6 respondents respectively. The Pearson chi-square statistics is 83.73 with an asymptotic significance p-value of 0.000. The result is significant if the p-value is less than or equal to the alpha value of 0.05. In this case, the p-value of 0.000 is less than the 0.005 alpha value, therefore we would reject the null hypothesis that asserts that the two variables age and coronavirus anxiety score (CAS) are independent of each other.

#### 4.1.3 Age and GAD score

**Table 3:** Age Distribution of Sample * Distribution GAD Cross tabulation.

| Independent Variable | GAD score | | | Total | $\chi^2$<br>(Pearson Chi-square) | P-value |
| --- | --- | --- | --- | --- | --- | --- |
| Age (Years) | 0-5 | >5-10 | >10-15 |  | 128.85 | .000 |
| 21-29 | 24 | 0 | 0 | 24 |  |  |
| 30-39 | 28 | 13 | 0 | 41 |  |  |
| 40-49 | 0 | 15 | 1 | 16 |  |  |
| 50-59 | 0 | 0 | 5 | 5 |  |  |
| 60-69 | 0 | 0 | 6 | 6 |  |  |
|  | 52 | 28 | 12 | 92 |  |  |

The table above presents the cross tabulation of generalized anxiety disorder (GAD) scores and age distribution of respondents. It could be observed that all 24 of 52 respondents identified with having a low GAD (0-5) are between the ages of 21-29 years of age while 28 respondents were 30- 39 years of age. 13 respondents of 28 respondents who have moderate GAD (>5-10) are between the ages of 30-39 while 15 respondents who have moderate GAD (>5-10) are in the age bracket of 40-49. A single respondent identified with high GAD (>10; 15) and is between the ages of 40-49. Also five res-spondents who suffered high GAD (>10; 15) falls within the 50-59 years of age. Similarly, 6 respondents who suffered high GAD (>10; 15) fall within the 60-69 years of age. The Pearson chi-square (128.85) shows a relationship between GAD scores and workplace risk. This is confirmed by its p-value of 0.000, which is less than the alpha value of 0.05. Therefore, we reject the null hypothesis that states that there is no relationship between GAD and age distribution and we accept the alternate hypothesis that there is a relationship between GAD and age distribution. All statistical tests were carried out at 5% level of significance.

#### 4.1.4 Profession and GAD Score

**Table 4:** distribution of profession against GAD score.

| Independent Variable | GAD score | | | $\chi^2$<br>(Pearson Chi-square) | p-value | Total |
| --- | --- | --- | --- | --- | --- | --- |
| Professions | 0-5 | >5-10 | >10-15 | 110.65 | .000 |  |
| Doctors | 15 | 10 | 3 |  |  | 28 |
| Nurses | 25 | 17 | 9 |  |  | 51 |
| Pharmacists | 5 | 1 | 0 |  |  | 6 |
| Med. Lab | 7 | 0 | 0 |  |  | 7 |
| Total | 52 | 28 | 12 |  |  | 92 |

The table above presents the cross tabulation of the various profession and their GAD scores. It could be observed that only 3 medical doctors had GAD scores >10, however, 9 nurses had GAD score of > 10. However, all of the Med. Lab technicians and 5 pharmacists out 6 scored between 0-5. The Pearson chi-square (110.65) shows a relationship between the various health professions (doctors, nurses, pharmacists, and medical laboratory technicians) and their GAD scores. This is confirmed by the p-values of 0.000, which is less than the alpha value of 0.05. Therefore, we reject the null hypothesis that states that there is no relationship between GAD and the various professions and we accept the alternate hypothesis that there is a relationship.

#### 4.1.5 Distribution of Profession

**Table 5:** Distribution of profession.

|  |  | Frequency | Percent | Valid Percent | Cumulative Percent |
| --- | --- | --- | --- | --- | --- |
| Valid | Doctors | 28 | 30.4 | 30.4 | 30.4 |
|  | Nurses | 51 | 55.4 | 55.4 | 85.9 |
|  | Pharmacists | 6 | 6.5 | 6.5 | 92.4 |
|  | Med. Lab | 7 | 7.6 | 7.6 | 100.0 |
|  | Total | 92 | 100.0 | 100.0 |  |

From the above it can be observed that 28 respondents, which are equivalent to 34% of the sample population, identified as doctors. Furthermore the data reveals that more than half of the respondents were nurses with 51 respondents identifying as nurses, which represents 55.4% of the sample population sample (92). More so, 6 respondents, which constitute 6.5%, identified as pharmacists. While 7 respondents, which represent 7.6% of the sample, population identified as med. Lab scientist.

#### 4.1.6 CAS and HCWs

**Table 6:**
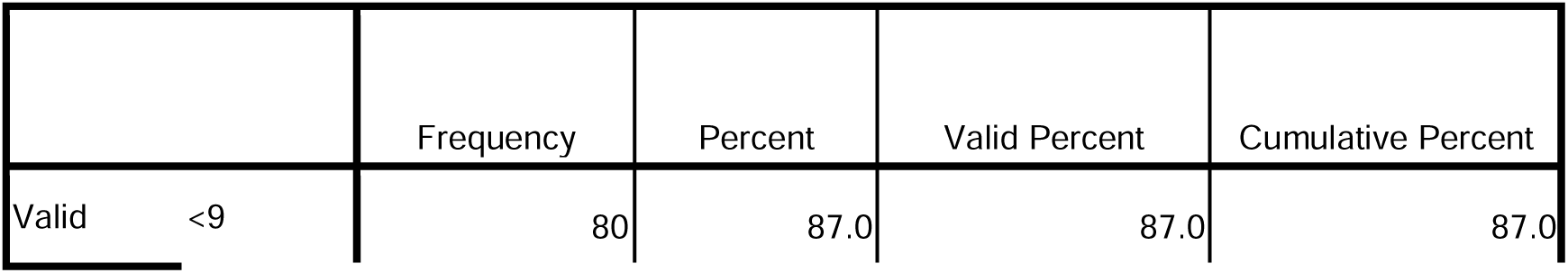
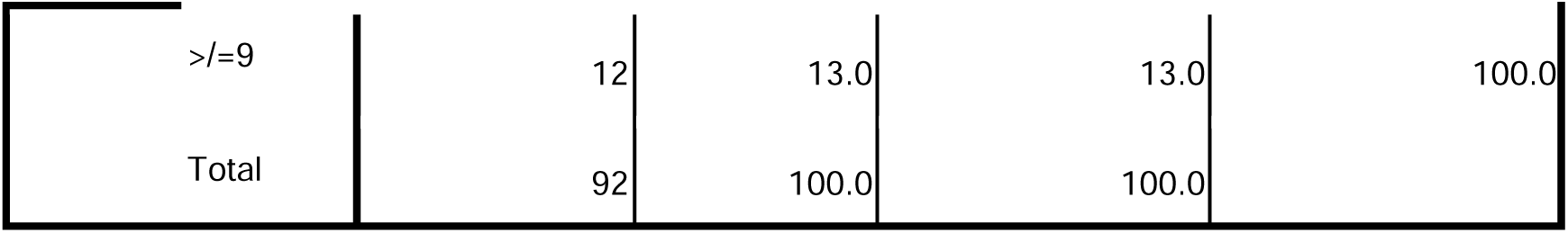
Distribution of CAS among the HCWs.

|  |  | Frequency | Percent | Valid Percent | Cumulative Percent |
| --- | --- | --- | --- | --- | --- |
| Valid | <9 | 80 | 87.0 | 87.0 | 87.0 |

|  |  |  |  |  |
| --- | --- | --- | --- | --- |
| >=9 | 12 | 13.0 | 13.0 | 100.0 |
| Total | 92 | 100.0 | 100.0 |  |

From the data above it can be observed that more than half of the respondents (80 respondents) which is equivalent to 87% of the sample population have a less severe CAS score of <9. More so, 12 respondents which constitute 13% seems to have severe coronavirus anxiety as revealed by their CAS score which was >=9.

#### 4.1.7 GAD Score and HCWs

**Table 7:** Distribution of GAD.

|  | Frequency | Percent | Valid Percent | Cumulative Percent |
| --- | --- | --- | --- | --- |
| Valid 0-5 | 52 | 56.5 | 56.5 | 56.5 |
| >5-10 | 28 | 30.4 | 30.4 | 87.0 |
| >10-15 | 12 | 13.0 | 13.0 | 100.0 |
| Total | 92 | 100.0 | 100.0 |  |

From the data above it can be observed that slightly above half of the respondents (52 respondents), which is equivalent to 56.5% of the sample population, have a low GAD score of between 0-5. However, 28 respondents which constitute 30.4% had a moderate GAD score of between >5-10. While 12 respondents which represented 13% of the sample population had high GAD score of between >10-15.

#### 4.1.8 Age Distribution of Sample

**Table 8:** Age Distribution of Sample.

|  | Frequency | Percent | Valid Percent | Cumulative Percent |
| --- | --- | --- | --- | --- |
| Valid 21-29 | 24 | 26.1 | 26.1 | 26.1 |
| 30-39 | 41 | 44.6 | 44.6 | 70.7 |
| 40-49 | 16 | 17.4 | 17.4 | 88.0 |
| 50-59 | 5 | 5.4 | 5.4 | 93.5 |
| 60-69 | 6 | 6.5 | 6.5 | 100.0 |
| Total | 92 | 100.0 | 100.0 |  |

From the data above it can be observed that 24 respondents which is equivalent to 26.1% of the sample population are within the age bracket 21-29 years. 41 respondents, which represent 44.6%, are within 30-39 years of age. Furthermore 16 respondents, which constitute 7.4%, are within 40-49 years of age, and 5 respondents, which correspond to 5.4%, fall in the age bracket 50- 59 years. Lastly 6 respondents which represents 6.5% of the sample population were 60 years and above.

#### 4.1.9 GAD level by Profession

**Table 9:** GAD by profession.

|  | Frequency | Percent | Valid Percent | Cumulative Percent |
| --- | --- | --- | --- | --- |
| Valid Doctors (0-5) | 15 | 16.3 | 16.3 | 16.3 |
| Doctors (>5-10) | 10 | 10.9 | 10.9 | 27.2 |
| Doctors >10-15) | 3 | 3.3 | 3.3 | 30.4 |
| Nurses (0-5) | 25 | 27.2 | 27.2 | 57.6 |
| Nurses (>5-10) | 17 | 18.5 | 18.5 | 76.1 |
| Nurses (>10-15) | 9 | 9.8 | 9.8 | 85.9 |
| Pharmacists (0-5) | 5 | 5.4 | 5.4 | 91.3 |
| Pharmacists >5-10 | 1 | 1.1 | 1.1 | 92.4 |
| Med.Lab (0-5) | 7 | 7.6 | 7.6 | 100.0 |
| Total | 92 | 100.0 | 100.0 |  |

From the table above, low GAD score is more prevalent among doctors with 15 respondents out of 28 respondents having GAD score of between 0-5 which represents 16.3% of the sample size, compared to 10 respondents having GAD score >5-10 which represents 10.9%, while 3 respondents which represents 3.3% had GAD score of between >10-15. Likewise for nurses, low GAD score was more prevalent with 25 respondents outs of 51 nurses which correspond to 27.2% of the sample population had GAD score of between 0-5, 17 respondents representing 18.5%, 9 respondents representing 9.8% of the sample population had GAD score of between >5-10 and GAD score of between >10-15 respectively. For the respondents who identified pharmacists, low GAD score was prevalent as 5 respondents out of 6 pharmacists representing 5.4% of the sample population had GAD score of between 0-5 and only 1 pharmacist GAD score of between >5-10 which is equivalent to 1.1& of the sample population.

#### 4.1.10 Gender and GAD score

**Table 10:** the relationship between gender and GAD score.

| Independent Variable | Sex | | | $\chi^2$<br>(Pearson) | p-value |
| --- | --- | --- | --- | --- | --- |
| GAD | Male | Female | Total | 34.243<sup>a</sup> | 0.000 |
| 0-5 | 30 | 22 | 22 |  |  |
| >5-10 | 0 | 28 | 28 |  |  |
| >10-15 | 0 | 12 | 12 |  |  |
|  | 30 | 62 | 62 |  |  |

The data above presents the cross tabulation of generalized anxiety disorder (GAD) scores and sex/gender of respondents. It could be observed that 30 out of 52 respondents identified with having a low GAD (0-5) are male while 22 respondents are female. Similarly, all 28 respondents who have moderate GAD (>5-10)) were female. Similarly, all 12 respondents who suffer high GAD (>10; 15) were female. The Pearson chi-square and likelihood ratio statistics shows a relationship between GAD score and sex/gender of respondents. This is confirmed by their p-values of 0.000 and 0.000 respectively, which is less than the alpha value of 0. 05. Therefore, we reject the null hypothesis that states that there is no relationship between GAD and sex/gender.

### 4.2 Work Place factors

#### 4.2.1 Work place risk and GAD

**Table 11:**
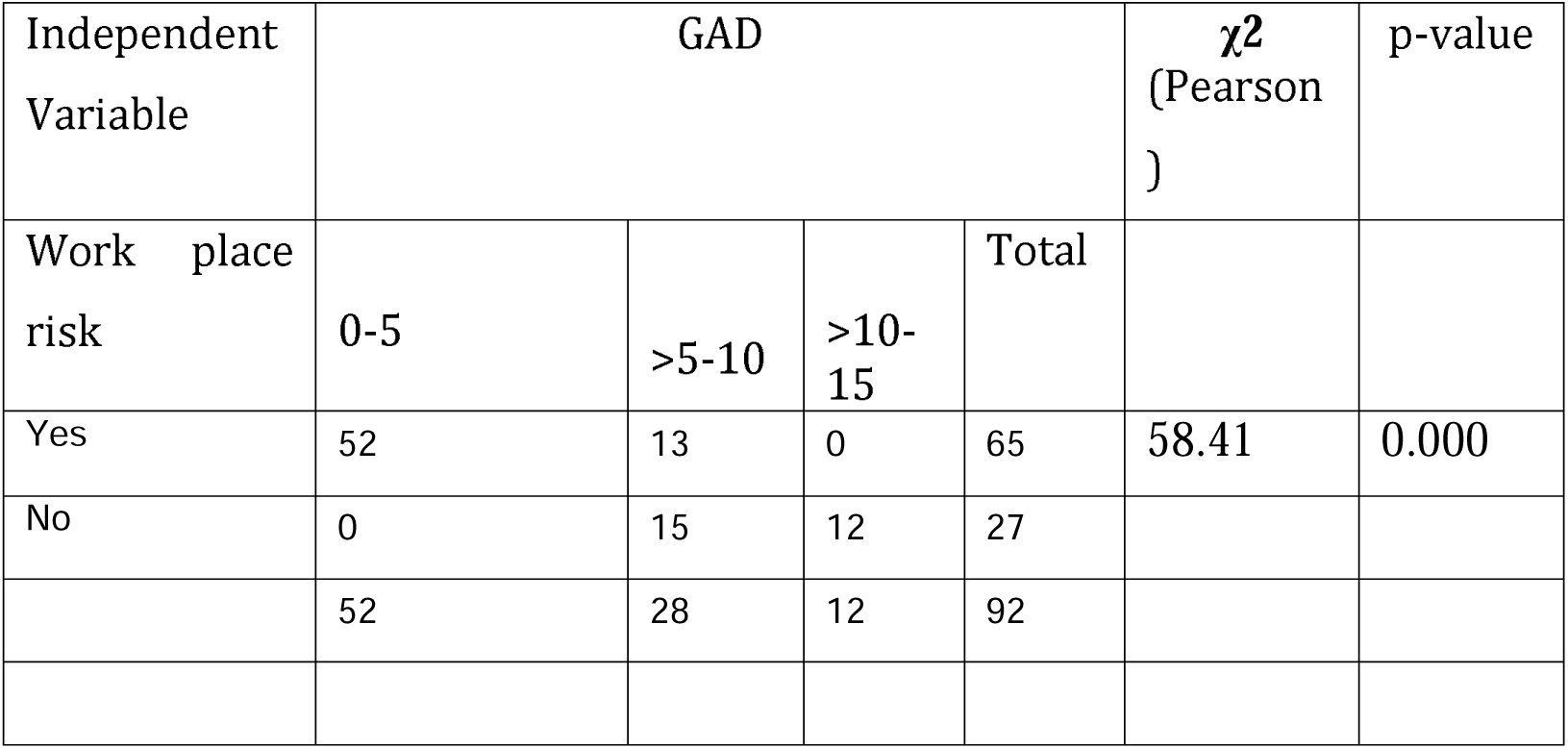
relationship between workplace risk and GAD.

| Independent Variable | GAD | | | | $\chi^2$<br>(Pearson) | p-value |
| --- | --- | --- | --- | --- | --- | --- |
| Work place risk | 0-5 | >5-10 | >10-15 | Total |  |  |
| Yes | 52 | 13 | 0 | 65 | 58.41 | 0.000 |
| No | 0 | 15 | 12 | 27 |  |  |
|  | 52 | 28 | 12 | 92 |  |  |

The table above presents the cross tabulation of generalized anxiety disorder (GAD) scores and workplace risk It could be observed that 50 out of 65 respondents identified with having a low GAD (0-5) believe they are exposed to risk at work, 13 respondents who have moderate GAD (>5- 10) believe they are exposed to risk at workplace risk while 15 respondents who have moderate GAD (>5-10) do not believe they are exposed to risk at work. Similarly, 12 respondents who suffer high GAD (>10; 15) do not believe there is risk at work. The Pearson chi-square (58.41) shows a relationship between GAD scores and workplace risk. This is confirmed by its p-value of 0.000, which is less than the alpha value of 0.05. Therefore, we reject the null hypothesis that states that there is no significant relationship between GAD and workplace and we accept the alternate hypothesis that there is a significant relationship. All statistical tests were carried out at 5% level of significance.

#### 4.2.2 GAD Score and PPE availability

**Table 12:** Table showing the relationship between GAD score and PPE.

| Independent Variable | Availability of PPE | | | $\chi^2$ (Pearson) | - alue |
| --- | --- | --- | --- | --- | --- |
| GAD | Yes | No | T tal | 13.63 | 0.001 |
| 0-5 | 52 | 0 | 52 |  |  |
| >5-10 | 28 | 0 | 28 |  |  |
| >10-15 | 10 | 2 | 12 |  |  |
|  | 90 | 2 | 92 |  |  |

The data above presents the cross tabulation of generalized anxiety disorder (GAD) scores and availability of PPE. It could be observed that 52 out of 52 respondents identified with having a low GAD (0-5) believe there is adequate availability of PPE. 28 respondents who have moderate GAD (>5-10) believe there is adequate availability of PPE while 10 respondents who have moderate GAD (>5-10) do not believe there is adequate availability of PPE at work, while 2 respondents who suffers high GAD (>10; 15) do not believe there is adequate availability of PPE at work. The Pearson chi-square (13.63) shows a relationship between GAD scores and availability of PPE. This is confirmed by the p-values of 0.001 that is less than the alpha value of 0. 05. Therefore, we reject the null hypothesis that states that there is no relationship between GAD and workplace and we accept the alternate hypothesis that there is a relationship.

#### 4.2.3 Previous Psychiatric Diagnosis and GAD

**Table 13:** Representation of Previous Psychiatric Diagnosis with GAD score.

| Independent variable | Availability of PPE | | | $\chi^2$<br>(Pearson ) | p-value |
| --- | --- | --- | --- | --- | --- |
| GAD | Yes | No | Total | 3.22 | 0.200 |
| 0-5 | 4 | 48 | 52 |  |  |
| >5-10 | 0 | 28 | 28 |  |  |
| >1 -15 | 0 | 12 | 12 |  |  |
|  | 4 | 88 | 92 |  |  |

The data above presents the cross tabulation of generalized anxiety disorder (GAD) scores and psychiatric condition of workers prior to coronavirus outbreak. It could be observed that 4 out of 52 respondents identified with having a low GAD (0-5) psychiatric condition prior to coronavirus outbreak while 48 out of 52 respondents identified with having a low GAD (0-5) did not psychiatric condition prior to coronavirus outbreak. Furthermore, all 28 respondents who have moderate GAD (>5-10) did not suffer psychiatric condition prior to coronavirus outbreak likewise all 12 respondents who suffers high GAD (>10; 15) did not suffer psychiatric condition prior to coronavirus outbreak. The Pearson chi-square (3.22) shows no relationship between GAD scores and psychiatric condition of workers prior to coronavirus outbreak. This is confirmed by the p-value of 0.200 that is greater than the alpha value of 0. 05. Therefore, we accept the null hypothesis that states that there is no relationship between GAD and psychiatric condition of workers and we reject the alternate hypothesis that there is no relationship. All statistical tests were carried out at 5% level of significance.

#### 4.2.4 CAS score and fear of COVID exposure

**Table 14:** Distribution of CAS score and fear of COVID-19 exposure.

| Independent variable | Fear of COVID-19 exposure | | | $\chi^2$<br>(Pearson ) | p-value |
| --- | --- | --- | --- | --- | --- |
| Distribution CAS | Yes | No | Total | 92.000<sup>a</sup> | 0.000 |
| <9 | 80 | 0 | 80 |  |  |
| >=9 | 0 | 12 | 12 |  |  |
|  | 80 | 12 | 92 |  |  |

The data above presents the cross tabulation of coronavirus anxiety score (CAS) scores and fear of contracting covid19 at work. It could be observed that only 80 respondents out of 80 respondents identified with a less severe CAS (<) have fear of contracting covid19 at work. Furthermore, all 12 respondents who have high CAS score (>=9) do not have fear of contracting covid19 at work. The Pearson chi-square is statistically significant at the 5% level with the asymptotic significance p-value of 0.000. In this case, the p-values of both statistics is less than the 0.05 alpha value, therefore we would reject the null hypothesis that asserts that the two variables coronavirus anxiety score (CAS) and fear of contracting covid19 at work is not significant and are independent of each other. In other words we accept the alternate hypothesis that states there is a relationship between coronavirus anxiety and fear of contracting covid19 at work.

#### 4.25 CAS and occupational support

**Table 15:** The distribution of CAS with regards to occupational support.

| Independent variable | Fear of COVID-19 exposure | | | $\chi^2$<br>(Pearson ) | p-value |
| --- | --- | --- | --- | --- | --- |
| Distribution CAS | Yes | No | Total | 4.082<sup>a</sup> | 0.043 |
| <9 | 21 | 59 | 80 |  |  |
| >=9 | 0 | 12 | 12 |  |  |
|  | 21 | 71 | 92 |  |  |

The data above presents the cross tabulation of coronavirus anxiety score (CAS) scores and occupational support. It could be observed that only 21 respondents out of 80 respondents identified with a less severe CAS (<) have occupational support. Furthermore, all 12 respondents who have high CAS score (>=9) do not have occupational support. The Pearson chi-square statistic is statistically significant at the 5% level with the asymptotic significance p-value of 0.043. The result is significant if the p-value is less than or equal to the alpha value of 0.05. In this case, the p-values of both statistics is less than the 0.05 alpha values, therefore we would reject the null hypothesis that asserts that the two variables coronavirus anxiety score (CAS) and occupational support have no relationship. In other words we accept the alternate hypothesis that states there is a relationship between coronavirus anxiety and occupational support.

## DISCUSSION

### 5.0 Main Findings and Comparison with literature

### 5.1 Age of HCWs and GAD

This study reported that being a young medical staff was negatively correlated with the development of GAD during the COVID-19 pandemic. In this study respondents within the age range of 21-49 years had mild (0-5) and moderate GAD scores, as opposed to those within the age bracket of 50-69 were all had moderate to severe GAD scores (>10 -15). This result contradicts the study by Lai et al (2020), which reported that being a young officer (as in < or = 30 years) is positively correlated to having increased levels of anxiety. The common knowledge that COVID-19 has a higher morbidity and mortality among the older population can explain the increased GAD levels among health workers aged 50 and above given their potentially high risk environment.

### 5.2 Gender of HCWs and GAD

Furthermore the female HCWs were more predisposed to developing GAD as opposed to their male counterparts. This is evidenced by the fact nurses had the most severe form of Corona phobia and GAD at 9.8%. Also the entire nursing staff is female. Again, out of the 62 respondents who were female, 22%, 28%, and 12% had mild, moderate, and severe GAD respectively. Their male counterparts all had mild anxiety. Liang et al (2020) also reported in their study in China that being female was positively related to elevated anxiety levels. However, study of anxiety levels amongst health professionals at an Ebola treatment unit in Liberia revealed anxiety especially among male staff (Li Li et al, 2020).

### 5.3 Previous psychiatric diagnosis and GAD

Retrospective studies in Toronto among HCWs during the SARS pandemic revealed that having a past history of mental illness increased ones risk of experiencing anxiety and other psychiatric symptoms (Lancee et al, 2008). However, in this study the relationship between previous psychiatric diagnosis prior to the outbreak of COVID-19 and GAD related to COVID-19 was insignificant. This contrasted with the study in Toronto.

### 5.4 High-risk station and GAD

During the MERS-CoV and SARS outbreaks, HCWs in units at high risk of infection present with more severe mental health outcomes compared with HCWs in units with low risk of infection (Bukhari et al, 2016). In my study, it was observed that 65 out of 92 respondents, who identified their workstation to be “high risk”, had some form of mild or moderate anxiety whereas all 27 respondents who considered their workstation not to be “high risk” had either moderate or severe anxiety.

### 5.5 Organizational support and GAD

In this study perceived organizational support was not necessarily a protective factor given that 21 out of 52 respondents who had mild GAD believed there would be organizational support if they came down with the virus. However, all 28 respondents who have moderate GAD and the 12 who have severe GAD do not believe there will be organizational support if they get infected. On the other hand a Canadian study reports organizational support as a protective factor among HCWs during the SARS outbreak (Fiksenbaum et al, 2006).

### 5.6 Confidence in PPE availability and GAD

Confidence in PPE availability was reported to not necessarily be a highly important protective factor given that 52 of the 90 respondents who believed that PPE was readily available suffered a mild GAD. This is supported by a study by Preti et al (2020) where it was revealed that availability of PPE only improves outcomes of stress and burnout but not necessarily the general mental state. Styra et al (2008) also comes to the same conclusion.

### 5.7 The GAD level amongst the different HCWs

The GAD level amongst doctors was 30.4% compared to nurses 55.4%. Furthermore, the prevalence among medical laboratory technicians and pharmacists were 7.6 % and 6.55 respectively. This revealed that anxiety is more prevalent in nurses. This corresponded to the prevalence values among doctors and nurses in Hongkong in 2003 during the SARS pandemic. In that study, anxiety was more prevalent in nurses (52.0%), than doctors (47.8%) during that pandemic (Poon et al, 2004). This further agreed with the findings by Matsuishi et al (2012) where they also reported that nurses had elevated anxiety during the H1N1 pandemic. A study narrated by Chakraborty (2020) of medical staff at a tertiary hospital during the COVID-19 pandemic supported the finding in the earlier study. Here, being female and working as a nurse was positively associated with increased anxiety. According to Kounou et al (2020) in Togo when grouped further, anxiety levels among HCWs was 25.8%, 22.6%, and 14.5% rates for mild, moderate, and severe anxiety respectively. This was high despite the relatively low infection rates in Togo. Kounou et al (2020) opined that these high rates could be accounted for by the poor quality of health care and lack of adequate PPE to deal with the COVID-19 pandemic in Togo. However the results in this study reveal that the anxiety levels among HCWs were 48.9%, 30.5%, and 20.7% for mild, moderate, and severe respectively. These figures are higher than that in Togo but reveal similar elevated levels of anxiety despite the fact that there are not many cases of COVID-19, and neither are the health facilities overwhelmed. A study of 231 HCWs in Mexico City using the Coronavirus Anxiety Scale (CAS) revealed that 30.3% of the participants were considered to have corona phobia having scored > or = 9 on the scale (Ignacio et al, 2020). However this study found that only 13.1% of HCWs were considered to have corona phobia.

### 5.1 Implications for practice

This study has revealed the impact of GAD on HCWs in a hospital in Ghana. Given the high prevalence recorded it is important to act quickly to handle the challenge of under funding and understaffing that have bedeviled both the delivery and access to mental health service. Age, female gender, being posted to high risk units and also being a nurse are characteristics that are positively linked to having elevated levels of GAD during the COVID-19 pandemic. This implies that people who are female, and nurses too are more likely to suffer morbidity due to COVID-19 related GAD. Furthermore female medical staff is less likely to be efficient in the execution of their duties as well as nurses since elevated anxiety levels can disturb the progress of work. Furthermore those with the aforementioned characteristics may present with restlessness, easy fatigability, irritability, muscle tension, and sleep disturbance. Given the crucial role of HCWs during a pandemic as front liners is vital and massive, making them susceptible to anxiety and stress. Therefore they are subject to complex emotional reactions and psychological distress that can impair their attention, cognitive functioning, and clinical decision-making (Pangioti et al, 2018). In the fight against COVID, “time is breath”. Thus if HCWs have impaired ability to take critical decisions for management or promptly identify symptom clusters then more life will be lost and there is greater chance of infection transmission. Prolonged unrecognized anxiety could lead to major psychiatric morbidity, exhaustion, and resignations from duties due to moral injury (Alharty et al, 2017). From this study it is also important that effort is made to make PPEs readily available to medical staff because this will curb GAD due to fear of exposure to infection. Again, it is important to consider the impact of perceived organizational support on the mental health of HCWs in light of the pandemic. Give that most of the doctors at FHH are locum doctors it is important to communicate support for their wellbeing and dump the notion that they are just “money making machines”. It is believed that this study will begin a conversation on how best to take care of the mental health of HCWs during this pandemic and beyond. It is hoped conversations on how to take care of the so-called “step-child” of health takes off. The findings of this study negate certain views in literature such as: that age less than or equal to 30 is positively related to GAD. Furthermore this study found that previous psychiatric diagnosis had no significant relationship with GAD since it had a Pearson chi-square value of 0.200. On the contrary various studies reported that previous psychiatric diagnosis was significantly related to GAD. However, this study supports the findings of previous studies reporting that organizational support, fear of exposure to COVID-19, and high risk work stations, and PPE access were significant in causing or preventing COVID-19 related GAD, given that in this study the P- values for the aforementioned variables was always less than the alpha value of 0.05.

### 5.3 Limitations

One challenge was obtaining a large sample for the study. The sample was relatively small and thus members of the group classified as health associates had to be added to the study population to boost the numbers. Furthermore this influenced the use of a consecutive sampling method that means that there might be bias towards a particular group of health care professions with regards to representation. Furthermore, since most of the doctors were locum doctors there was a difficulty in determining of their fear of COVID -19 exposure was tied their part time practice at FHH or if it was related to their practice at bigger hospitals that may have more patients with the infection.

## 6.0 CONCLUSION AND RECOMMENDATIONS

### 6.1 Conclusion

COVID-19 related GAD is a challenge amongst HCWs especially nurses in FHH. The GAD level among nurses was 55.4%, and for doctors it was 30.4%. The GAD level among medical laboratory technicians and pharmacists were 7.6% and 6.5% respectively. Prevalence of COVID-19 related GAD among health care workers in FHH. Furthermore being between 50-69 years is a significant risk factor for developing GAD during the COVID-19 pandemic in Ghana. However, only 13.1% of the HCWs were considered to have Corona phobia. Perception of workplace as being “high risk” was positively correlated with mild to moderate forms of anxiety. However, perception of organizational support as being guaranteed in the case one succumbed to the virus and confidence in PPE availability were not reported to be strong protective factors against GAD among HCWs.

### 6.2 Recommendations

**Based on the findings of this study the following are recommendations to be considered for policy and practice:**

1. Sensitization campaigns carried out by the Medical Director with liason with Ghana Health Service to help doctors identify symptoms of anxiety and encourage them to seek help.
2. The establishment of various hotlines and other mental health and psychosocial services (MHPSS) by the Ministry of health for HCWs to reach out for help once they start to experience certain symptoms of anxiety.
3. Furthermore the formation of routine support processes such as peer support programs should be encouraged among healthcare staff especially nurses.
4. Establishing and encouraging the use of online psychological counseling services as well as online psychological self-help intervention systems including online cognitive behavioral therapy for anxiety.
5. Communication and regular reminders about the existence of hotlines for HCWs.
6. The hospital management team should make extra effort to build HCWs confidence in perceived organizational support in case they succumb to the virus.
7. The hospital management should encourage adherence of COVID-19 prevention protocols among patients and staff alike and ensure that PPEs are readily available to protect HCWs especially those between the ages of 50-69.

## Supporting information

IRB Approval

## Data Availability

All data produced in the present work are contained in the manuscript

## INFORMED CONSENT FORM

### General Information about Research

This research is in fulfillment of the compulsory dissertation required of final year medical students at the said medical school. The aim of this study is to determine the prevalence of COVID-19 induced/related anxiety among health care professionals with the ultimate goal of encouraging health facilities/government to establish or fund several mental health and psychosocial services for health care professionals such as psychological hotlines during this pandemic. It is also hoped that these services will remain post pandemic to also cater to other mental health challenges health care professionals may face. In this study, participants are kept anonymous and confidentiality is guaranteed. The questionnaire that follows has been designed to take a maximum of 5 minutes to complete . Besides the socio-demographic aspect, this questionnaire uses both the Generalized Anxiety Scores and the Coronavirus Anxiety scores to measure anxiety levels in Health Care Workers.

### Possible Risks

There are no risks to participants but rather benefits to the health care profession.

### Possible Benefits

This study will promote the establishment and strengthening of mental health services for health care workers at Family Health Hospital,and by extension health facilities in Accra.

### Confidentiality

We will protect information about you and the questionnaires are anonymous.

### Compensation

Unfortunately, there is no compensation for participating in this study. The Principal Investigator deeply regrets this.

### Contacts for additional information

To obtain answers to pertinent questions about the research and whom to contact in case of research-related injury kindly contact the Principal Investigator, Ohakpougwu Chukwuebuka Emmanuel (0249386997), or His Supervisor Ms. Phaedra Yamson (0204519492).

## VOLUNTEER AGREEMENT

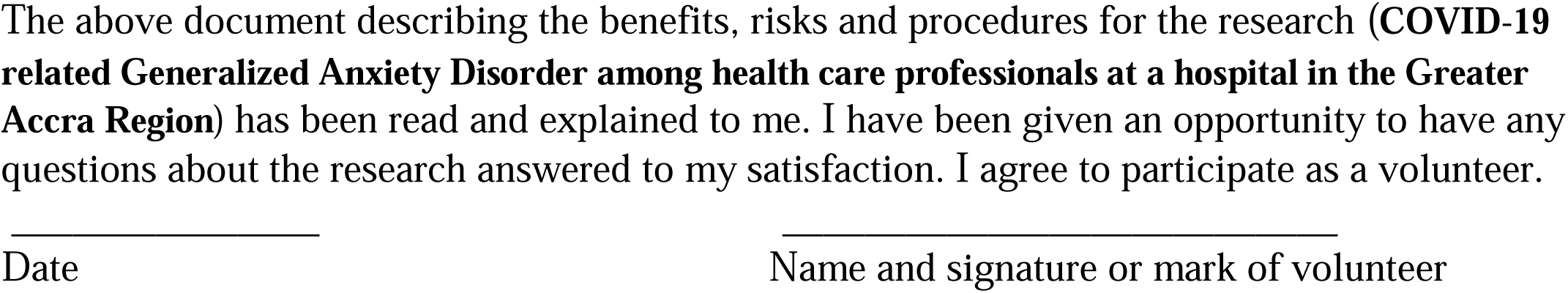

## QUESTIONNAIRE

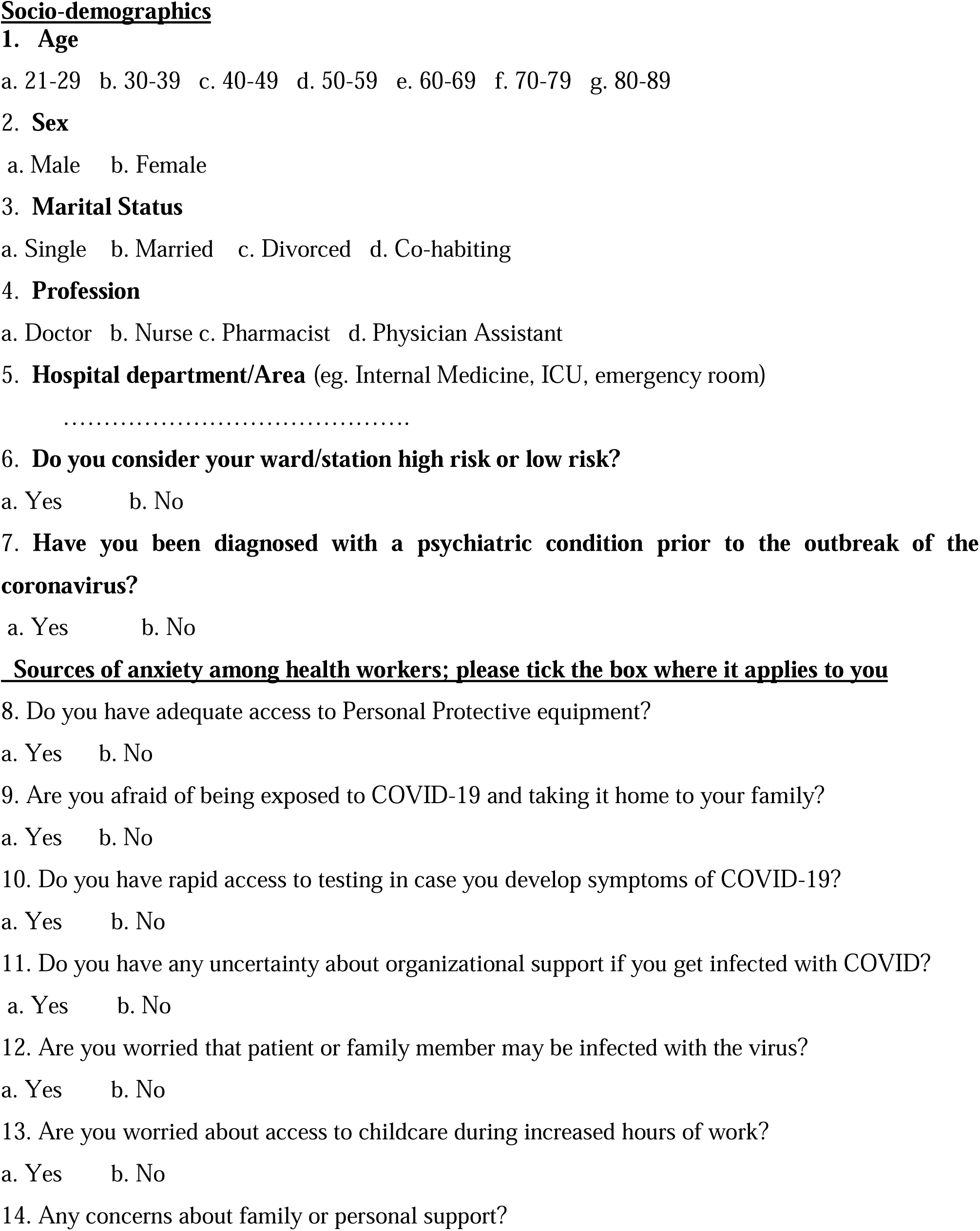
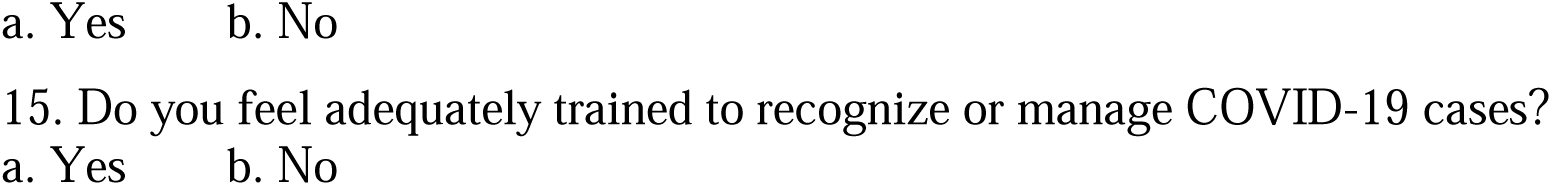

## PLEASE TICK THE OPTION THAT APPLIES BELOW BUT DO NOT SUM THE SCORE

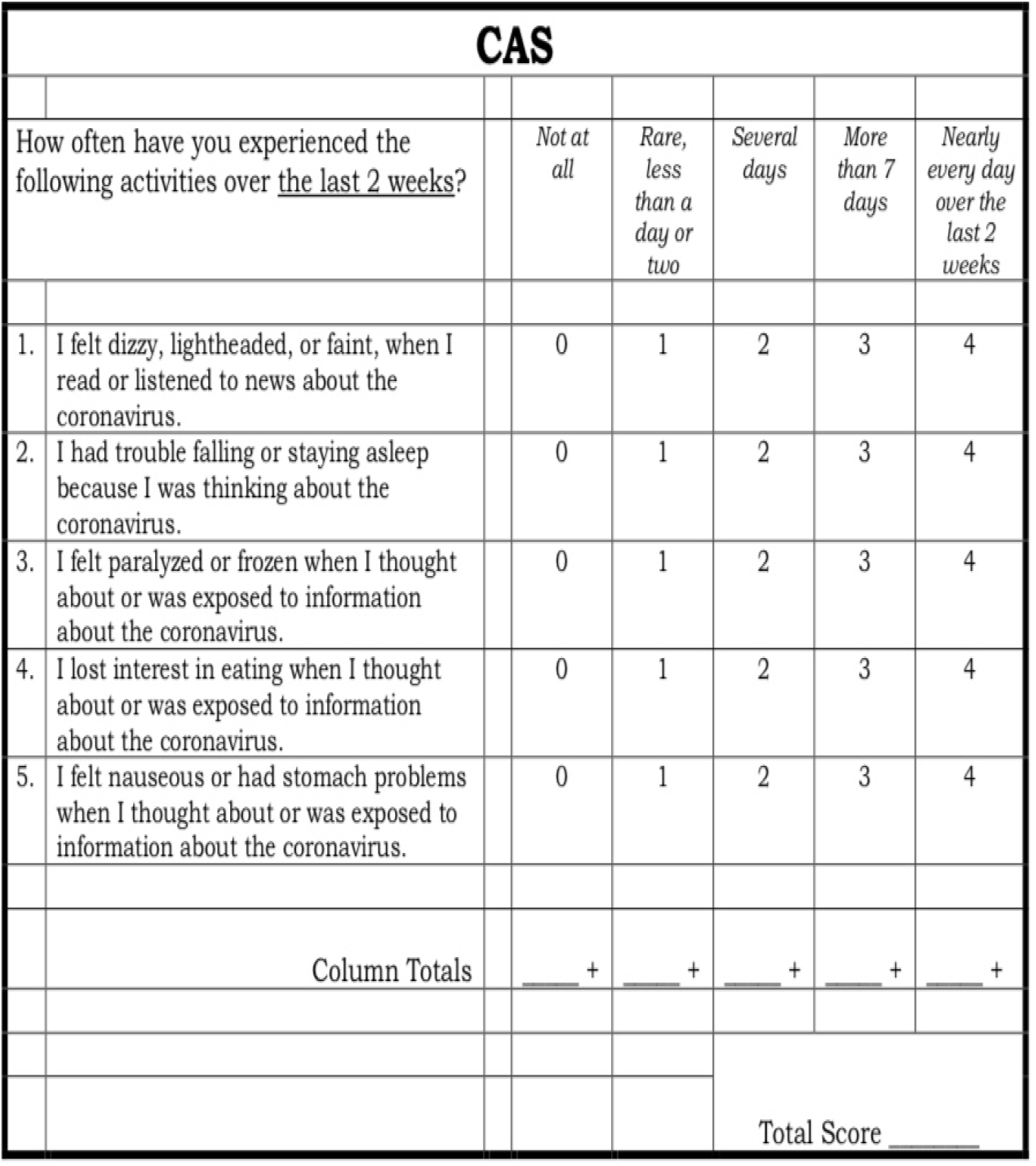

## TICK THE OPTION THAT APPLIES BELOW BUT DO NOT SUM THE SCORE

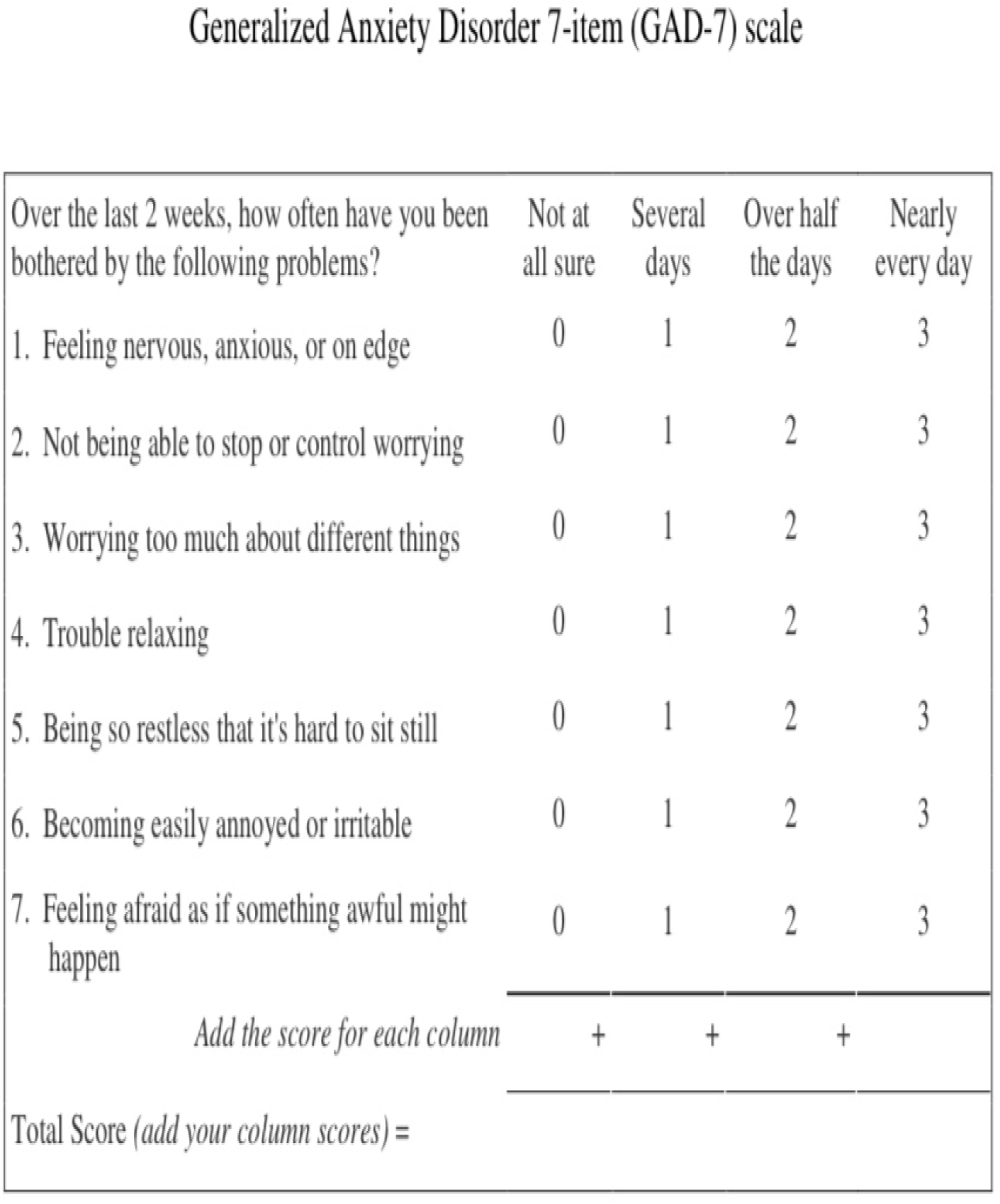

